# Clinical equipoise and patient preferences for DOAC resumption after high-risk endoscopy: implications for a randomized trial

**DOI:** 10.64898/2026.08.14.26360466

**Authors:** Zachary L. Smith, B. Joseph Elmunzer, Nauzer Forbes, Christian T. Ruff, Mellanie T. Hills, Denise M. Scholtens

## Abstract

The optimal timing of resuming direct oral anticoagulants (DOACs) after high-risk endoscopic procedures remains uncertain. Existing recommendations are largely based on expert opinion, resulting in substantial practice variability. Understanding clinician equipoise and patient preferences is essential to inform the design of randomized trials, including the proposed Resumption of Direct Oral Anticoagulants After High-Risk Endoscopy (RESUME) trial.

**Methods:** We conducted parallel, cross-sectional surveys of practicing endoscopists and patients with atrial fibrillation using electronic questionnaires administered via Qualtrics. The endoscopist survey, distributed through the American Society for Gastrointestinal Endoscopy, assessed practice patterns, acceptability of early (postoperative day [POD] +1), intermediate (POD +3), and late (POD +5) resumption strategies, and perceptions of clinical equipoise. The patient survey, distributed through two advocacy organizations, evaluated awareness of evidence gaps and prioritization of bleeding versus thromboembolic risk.

**Results:** A total of 201 endoscopists and 477 patients (92.5% taking a DOAC) were included. Endoscopists demonstrated wide variability in preferred timing of DOAC resumption after a standardized high-risk mucosal resection vignette, ranging from same-day resumption to delays beyond five days. POD +2 was the most commonly selected strategy, and most respondents rated more than one proposed RESUME trial arm as acceptable. Nearly all endoscopists (98.9%) rated a randomized trial to determine optimal timing as important. Patient preferences regarding bleeding versus stroke risk were heterogeneous and symmetrically distributed around the neutral response on a five-point ordinal scale. Preferences did not differ by prior stroke or transient ischemic attack, prior major bleeding, age, sex, or geographic region. More than half of patients reported confidence that clear guidance exists regarding DOAC resumption, despite the absence of high-quality randomized evidence informing postprocedural timing.

**Conclusions:** Both clinicians and patients demonstrate substantial variability and clinical uncertainty regarding optimal timing of DOAC resumption after high-risk endoscopy, supporting the ethical justification, feasibility, and relevance of the proposed RESUME trial.

## Background

Direct oral anticoagulants (DOACs) have become the most widely used medications for conditions which require the mitigation of thromboembolic risk, rapidly surpassing warfarin since their introduction to market^1-3^. A recent study of 17.3 million American adults with atrial fibrillation (AF) leveraging the Medical Expenditure Panel Survey reported that DOAC prevalence rose from 52.1% in 2016 to 81.6% in 2021^4^., with similar patterns also observed in Europe^5, 6^. As their use has increased, gastroenterologists are increasingly required to make periprocedural anticoagulation decisions for patients undergoing endoscopic procedures with a high risk of delayed bleeding.

Current guidelines provide general recommendations for periprocedural anticoagulation management but offer limited evidence to guide DOAC resumption following high-risk endoscopic interventions, including large/complex polyp resection, endoscopic retrograde cholangiopancreatography (ERCP) with sphincterotomy, variceal band ligation, transmural drainage procedures, and more^7-14^. Existing data, limited by mostly single arm observational studies, suggest that rates of delayed bleeding after high-risk endoscopic procedures can exceed 15-20%^14-19^. Therefore, existing guideline recommendations are largely based on low quality evidence, pharmacokinetic considerations, and expert opinion, contributing to substantial variability in clinical practice.

Decisions regarding anticoagulant resumption are thus inherently preference-sensitive, requiring clinicians to weigh competing risks and incorporate patient values, often with uncertainty. However, patient perspectives have rarely been incorporated into studies or guidelines addressing periprocedural anticoagulation management. Similarly, as clinician equipoise is essential for randomized trials, the extent to which endoscopists accept alternative resumption strategies has not been systematically evaluated.

The proposed RESUME trial will be a pragmatic, randomized trial comparing early, intermediate, and late DOAC resumption strategies following high-risk endoscopic procedures. RESUME aims to evaluate resumption strategies that are already in widespread use and considered acceptable by both clinicians and patients. To ensure ethical justification, feasibility, and relevance, it is critical to establish that both clinicians and patients demonstrate sufficient uncertainty and equipoise regarding various resumption timing. The aim of this study was to survey practicing endoscopists and patients with atrial fibrillation to assess current practice patterns, perceptions of evidence gaps, and prioritization of bleeding versus thromboembolic risk. These findings were intended to directly inform the design and conduct of the RESUME trial.

## Methods

### Study design and oversight

This was a cross-sectional, survey-based study of both attending gastroenterologists and patients with atrial fibrillation. The surveys were developed by the investigators to assess real-world practice patterns, clinical equipoise, and stakeholder priorities related to balancing bleeding and thromboembolic risk. Both surveys were administered electronically using the Qualtrics platform (Qualtrics, Provo, UT). Participation was voluntary and anonymous. Respondents were informed that the surveys were intended to inform clinical trial design. No incentives were provided for participation.

The clinician and patient surveys were reviewed and registered by the Medical College of Wisconsin Institutional Review Board (PRO00056619 and PRO00057092). Electronic informed consent was obtained using an informational letter prior to participation. No protected health information was collected.

### Study Development and Dissemination

Both survey instruments were developed through structured discussions among the investigative team in parallel with a focused review of the medical literature to identify areas of uncertainty in periprocedural anticoagulation management. The endoscopist survey was initially drafted by a single investigator (ZLS) and subsequently refined through iterative review and pilot testing by other investigators (BJE, CTR, DMS) to establish face and content validity and to improve clarity, question order, and ease of administration. Revisions were made to minimize ambiguity and to ensure that response options reflected real-world clinical practice.

The patient survey was developed by a single investigator (ZLS) in collaboration with a representative from StopAFib.org with extensive experience in qualitative survey research (MTH). The survey was developed iteratively to ensure readability for a lay audience. Survey items were written to minimize technical terminology and to present clinical tradeoffs in clear, patient-centered language. The instrument was reviewed by other investigators to improve clarity and comprehension prior to distribution.

The endoscopist survey was distributed electronically through the American Society for Gastrointestinal Endoscopy (ASGE) to attending gastroenterologists. Investigators applied for access to the ASGE membership email distribution list through the standard organizational process. The survey instrument was reviewed by the ASGE Research Committee and approved for distribution. The patient survey was distributed electronically via email through Arrhythmia Alliance and StopAFib.org. Survey content

### Endoscopist Survey

The endoscopist survey was designed to assess current practice patterns, perceptions of clinical equipoise, and acceptability of proposed DOAC resumption strategies following high-risk endoscopic procedures. The survey included a hypothetical clinical vignette describing a 74-year-old patient with nonvalvular atrial fibrillation undergoing conventional endoscopic mucosal resection (EMR) of a 35-mm sessile right-sided colorectal polyp without clip closure. Respondents were asked to select the single best timing for DOAC resumption in this scenario. Response options included: 0 days (resume same day), 1 day, 2 days, 3 days, 4 days, 5 days, more than 5 days, I am unsure, and defer to cardiology or the prescribing physician. Subsequent questions assessed the acceptability of three proposed RESUME trial arms—early (postoperative day [POD] +1), intermediate (POD +3), and late (POD +5) resumption—using a four-point Likert scale ranging from “not acceptable” to “strongly prefer.” Additional items assessed the perceived importance of a randomized trial to guide DOAC resumption, willingness to consider alternative resumption strategies, and the relative severity of bleeding- and stroke-related outcomes using a rank-order format. Demographic and practice characteristics collected included age, gender, years in independent practice, geographic region of practice, practice setting, annual ERCP volume, and annual volume of large polyp resections (≥20 mm EMR/ESD). The full endoscopist survey instrument is provided in the **Supplementary Material**.

### Patient survey

The patient survey was designed to assess patient awareness of evidence gaps, perceptions of bleeding and stroke risk, and prioritization of outcomes following high-risk gastrointestinal procedures. The survey was developed in collaboration with a patient advocacy organization (StopAFib.org) to ensure clarity and accessibility. Collected demographic variables included age group, gender, and continent of residence. Clinical history items assessed current or prior use of DOAC therapy, prior stroke or transient ischemic attack (TIA), prior major bleeding events, and prior gastrointestinal procedures. Patient awareness of existing evidence was assessed using a confidence-based question regarding the existence of clear guidance on DOAC resumption after high-risk procedures. Outcome prioritization was assessed using two complementary approaches: (1) a forced-choice question asking respondents to select which outcome concerned them most (major bleeding versus stroke with long-term disability), and (2) a five-point ordinal scale assessing relative concern for bleeding versus stroke. The full patient survey instrument is provided in the **Supplementary Material**.

### Survey Participants and Analysis Populations

The endoscopist survey was distributed via email through the ASGE to practicing attending gastroenterologists. It was not distributed to trainee or non-physician members. The patient survey was distributed via email through Arrhythmia Alliance and StopAfib.org, international patient advocacy organizations serving individuals with atrial fibrillation. Participation was open to adults >18 years of age with or without prior procedural experience. Respondents were not required to be currently taking a DOAC to participate. For the endoscopist survey, all respondents who provided evaluable responses to acceptability or prioritization questions were included in analyses relevant to those outcomes. For the patient survey, the analysis population included all respondents who completed at least one outcome prioritization question (forced-choice or ordinal scale). No imputation was performed for missing data. As a result, denominators varied across analyses based on item-specific response completion.

### Statistical Analysis

Survey responses were summarized using descriptive statistics. Categorical variables are reported as counts and percentages. For the patient survey, the analysis population included all respondents who completed at least one outcome prioritization question. No imputation was performed for missing data; analyses were conducted using available responses for each survey item, and denominators therefore varied across analyses. Patient prioritization of bleeding versus thromboembolic risk was assessed using both a forced-choice question and a five-point ordinal scale. Ordinal responses were analyzed using nonparametric methods. Comparisons between two groups were performed using the Wilcoxon rank-sum test, and comparisons across more than two groups were performed using the Kruskal–Wallis test. Associations between categorical variables were evaluated using Pearson’s chi-squared test; Fisher’s exact test was used in cases of small expected cell counts.

Subgroup analyses evaluating associations between clinician characteristics and acceptability of proposed DOAC resumption strategies were considered exploratory. No adjustment was made for multiple comparisons. All statistical tests were two-sided, and p values <0.05 were considered nominally significant. Statistical analyses were performed using R (R Foundation for Statistical Computing, Vienna, Austria).

## Results

### Survey respondents and baseline characteristics

#### Endoscopist survey

A total of 201 endoscopists completed the clinician survey. Baseline characteristics are present in **Table 1**. Respondents represented a wide range of years in independent practice, practice settings, and procedural experience, including ERCP and advanced polyp resection. Most respondents practiced in academic or private practice settings. Nearly all respondents (98.9%) indicated that a randomized trial evaluating DOAC resumption after high-risk endoscopic procedures was important, with the majority rating such a trial as very or extremely important **(Supplemental Table 1)**.

**Table 1:** Baseline Characteristics of Surveyed Gastroenterologists.

| **Age (N=200)** | **Endoscopists (N=201)** |
| --- | --- |
| **Under 35** | 17 (9.9%) |
| **35-44** | 53 (31%) |
| **45-54** | 30 (18%) |
| **55-64** | 37 (22%) |
| **65 or older** | 33 (19%) |
| **Unknown** | 30 |
| **Gender (N=199)** |  |
| **Male** | 142 (84%) |
| **Female** | 26 (15%) |
| **Unknown** | 31 |
| **Years in independent practice (N=200)** |  |
| **0-5 years** | 49 (29%) |
| **6-10 years** | 22 (13%) |
| **11-15 years** | 14 (8.2%) |
| **16-20 years** | 20 (12%) |
| **21 or more years** | 65 (38%) |
| **Unknown** | 30 |
| **Primary practice setting (N=201)** |  |
| **Academic medical center** | 61 (37%) |
| **Private practice** | 57 (34%) |
| **Hybrid + VA + Other** | 49 (29%) |
| **Unknown** | 34 |
| **ERCPs performed per year (average) (N=200)** |  |
| **0-25 / Do not perform** | 94 (55%) |
| **26+** | 76 (44%) |
| **Unknown** | 30 |
| **Large polyp resections (EMR/ESD) performed per year (average) (N=200)** |  |
| **0-10 / Do not perform** | 32 (22%) |
| **11+** | 110 (77%) |
| **Unknown** | 58 |
| Denominators for each variable where <100% answered are noted. |  |

#### Patient survey

A total of 533 patients initiated the patient survey. Of these, 477 respondents met criteria for inclusion in the analysis population by completing at least one outcome prioritization question. Baseline characteristics of patient respondents are shown in **Table 2**. Most respondents were older adults, with a predominance of women. The majority resided in North America, with additional representation from Europe and other regions. Most (92.6%) reported current use of a direct oral anticoagulant. A history of prior stroke or transient ischemic attack was reported by 64 respondents (13.4%), and 38 respondents (8.0%) reported a prior major bleeding event.

**Table 2:**
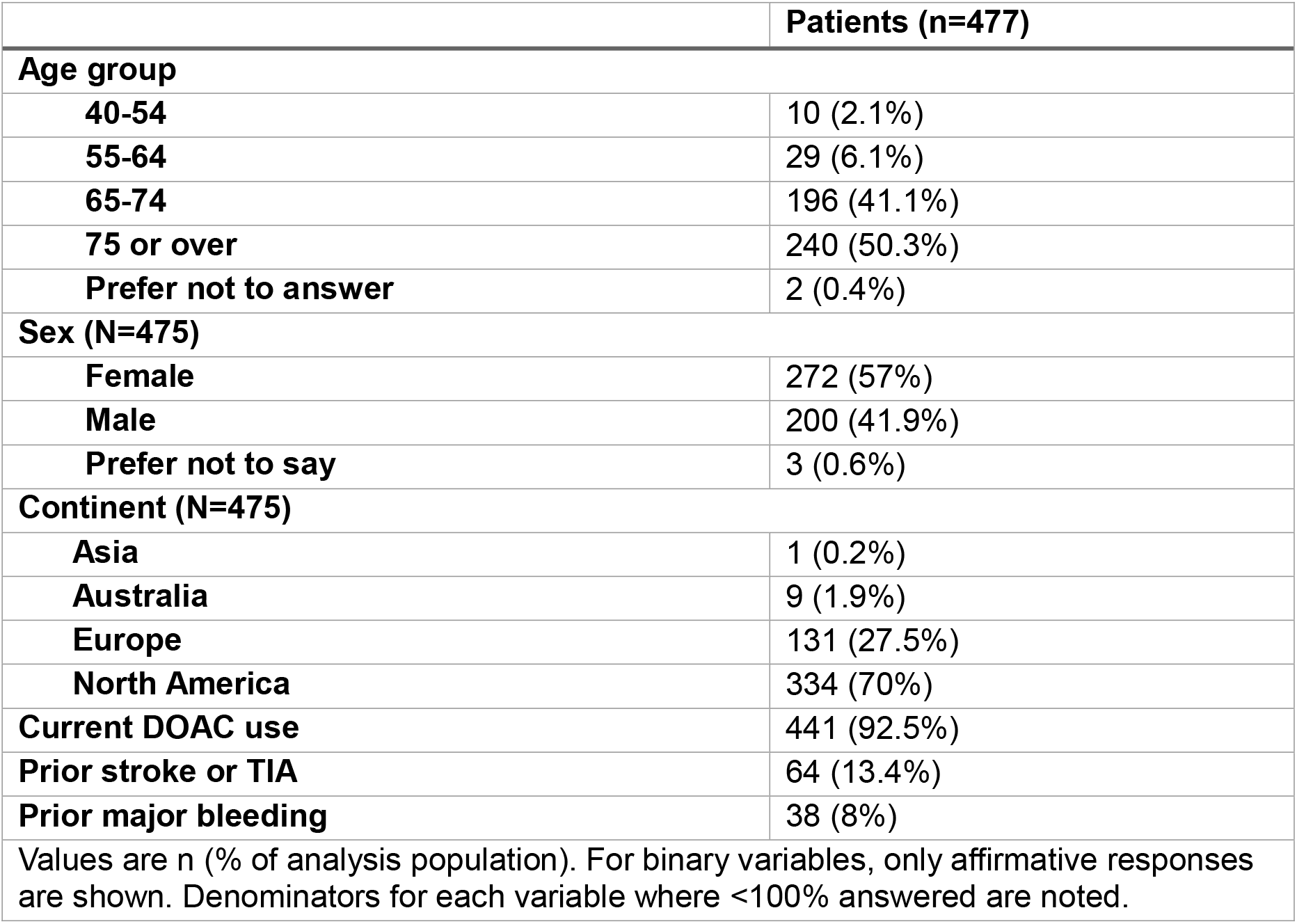
Baseline characteristics of surveyed patients.

### Endoscopist practice patterns and clinical equipoise

Endoscopists demonstrated substantial variability in their approach to DOAC resumption following a standardized clinical vignette involving high-risk colorectal polyp resection **(Figure 1)**. While postoperative day (POD) +2 was the most common resumption time, responses ranged from same-day resumption to delays exceeding five days, highlighting the absence of a dominant practice pattern **(Supplemental Table 2)**.

**Figure 1:**
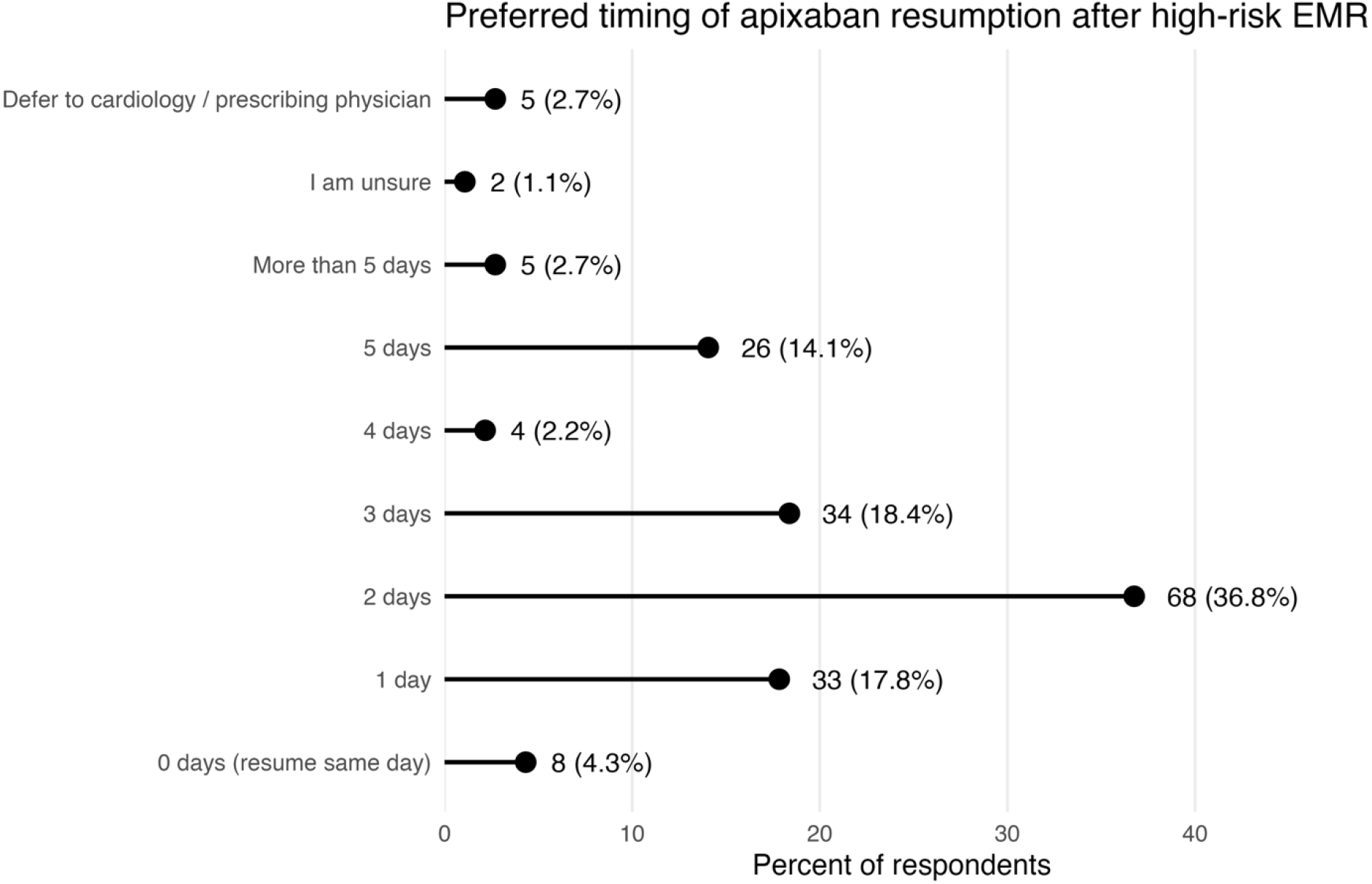
Distribution of responses regarding time to resuming apixaban after hypothetical conventional EMR clinical vignette

When asked about the acceptability of the three proposed RESUME trial arms (POD +1, POD +3, and POD +5), most respondents endorsed more than one strategy as acceptable. POD +3 was acceptable to the largest proportion of respondents (94.5%), while POD +1 (72.3%) and POD +5 (70.7%) were also considered acceptable by substantial majorities. Only a small minority of respondents (2.5%) found both early and late resumption strategies unacceptable **(Table 3, Supplemental Table 3, Supplemental Table 4)**.

**Table 3:**
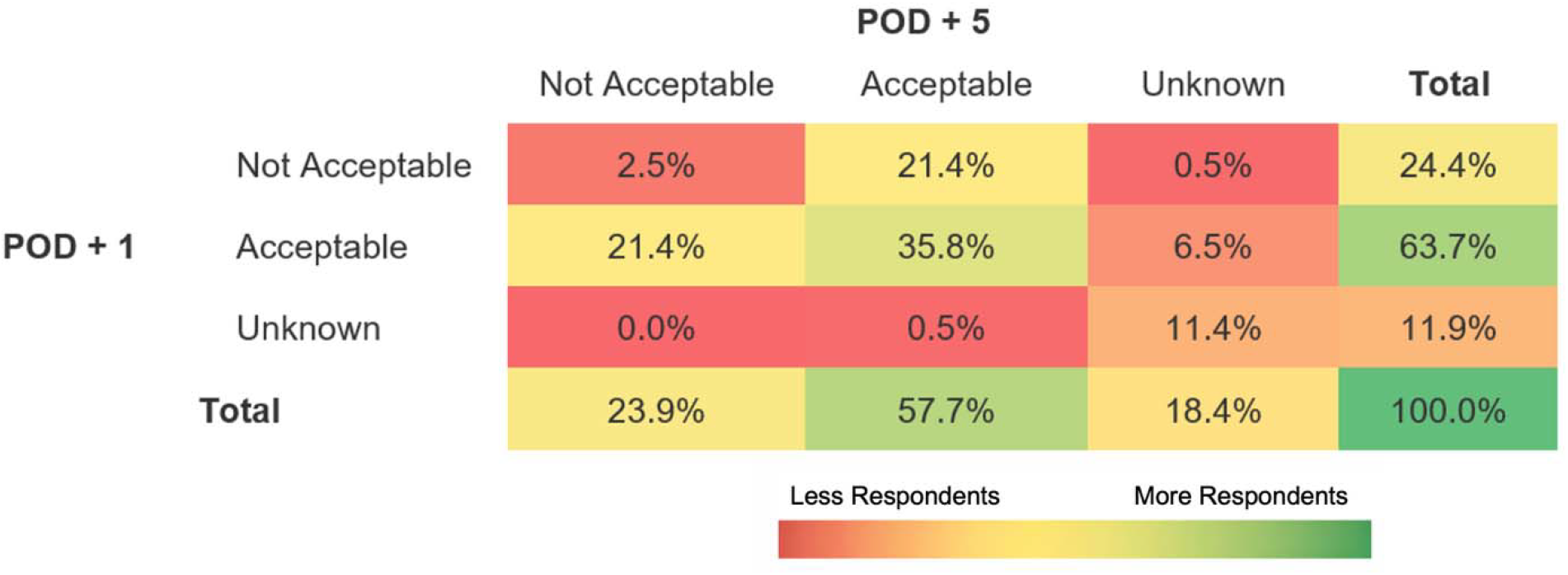
Proportion of respondents finding postoperative day (POD) +1 and +5 acceptable or unacceptable for a hypothetical conventional mucosal resection of a large colorectal polyp.

|  |  | POD + 5 |  |  | Total |
| --- | --- | --- | --- | --- | --- |
|  |  | Not Acceptable | Acceptable | Unknown |  |
| POD + 1 | Not Acceptable | 2.5% | 21.4% | 0.5% | 24.4% |
|  | Acceptable | 21.4% | 35.8% | 6.5% | 63.7% |
|  | Unknown | 0.0% | 0.5% | 11.4% | 11.9% |
| Total |  | 23.9% | 57.7% | 18.4% | 100.0% |

Less Respondents More Respondents

**Table 4:**
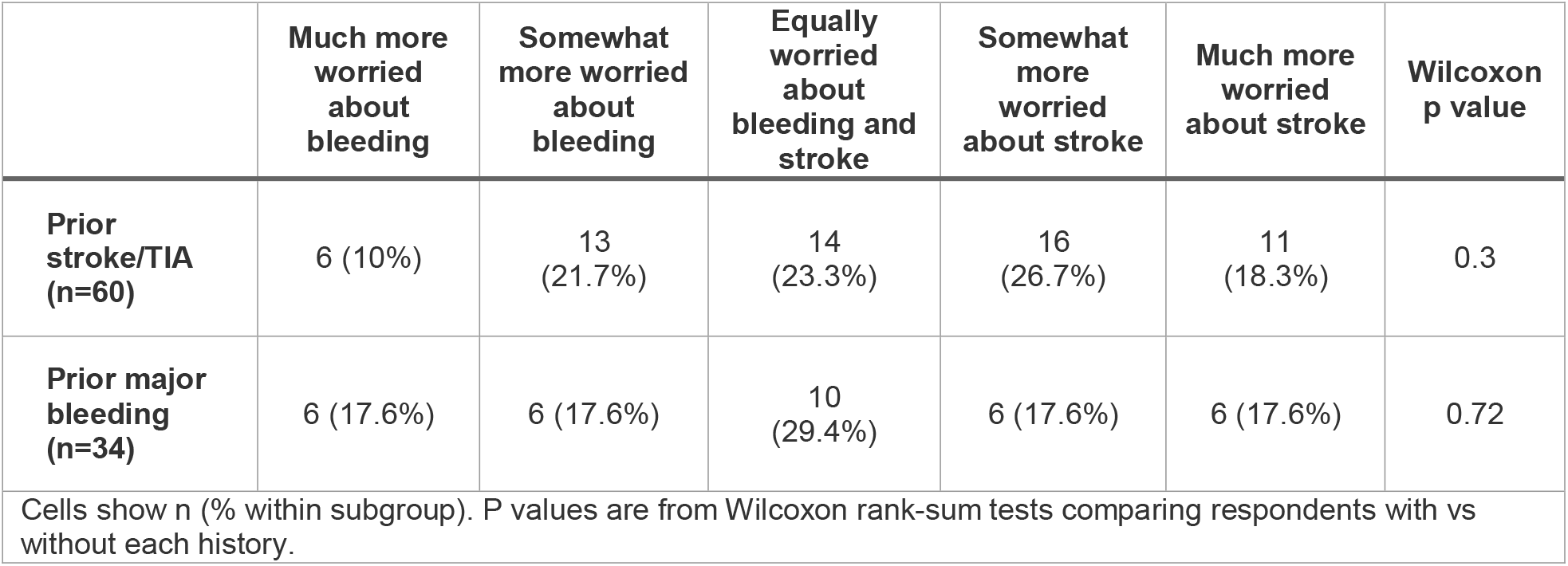
Outcome prioritization in patients with prior stroke and/or major bleeding.

|  | Much more worried about bleeding | Somewhat more worried about bleeding | Equally worried about bleeding and stroke | Somewhat more worried about stroke | Much more worried about stroke | Wilcoxon p value |
| --- | --- | --- | --- | --- | --- | --- |
| **Prior stroke/TIA (n=60)** | 6 (10%) | 13 (21.7%) | 14 (23.3%) | 16 (26.7%) | 11 (18.3%) | 0.3 |
| **Prior major bleeding (n=34)** | 6 (17.6%) | 6 (17.6%) | 10 (29.4%) | 6 (17.6%) | 6 (17.6%) | 0.72 |

Cells show n (% within subgroup). P values are from Wilcoxon rank-sum tests comparing respondents with vs without each history.

When ranking the severity of potential clinical outcomes, respondents consistently rated death from stroke as the most severe outcome, followed by death from delayed bleeding, stroke with permanent neurologic deficits, delayed bleeding requiring intervention, transient ischemic attack, and self-limited delayed bleeding **(Supplemental Table 5)**.

### Associations between clinician characteristics and acceptability of resumption strategies

Exploratory subgroup analyses evaluated associations between clinician characteristics and acceptability of early (POD +1) and late (POD +5) DOAC resumption strategies. These analyses included years in practice, practice setting, annual ERCP volume, and annual large polyp resection volume. Across subgroups, acceptability of POD +1 and POD +5 was generally consistent, and no robust or consistent associations were observed. While isolated nominal differences were observed in exploratory analyses, these findings were not consistent across subgroup comparisons and were not adjusted for multiple testing. Overall, acceptability of both early and late resumption strategies appeared broadly similar across clinician characteristics **(Supplemental Tables 6-13)**.

### Patient awareness of evidence gaps

Patient awareness of evidence guiding DOAC resumption after high-risk procedures was variable. Slightly more than half of respondents (54.6%) reported being very or somewhat confident that clear scientific evidence or medical guidance exists regarding the timing of DOAC resumption **(Supplemental Table 14)**. However, a substantial proportion of respondents expressed uncertainty or lack of confidence.

### Patient prioritization of bleeding versus thromboembolic risk

Patient preferences regarding the competing risks of delayed gastrointestinal bleeding and stroke were assessed using both a forced-choice question and a five-point ordinal scale. Forced-choice responses were divided between prioritizing bleeding-related complications and stroke-related complications, without a dominant preference emerging. On the ordinal scale, responses demonstrated a symmetric distribution centered around the neutral response (i.e. equipoise), with the largest proportion of respondents reporting equal concern for bleeding and stroke **(Figure 2, Supplemental Table 15)**.

**Figure 2:**
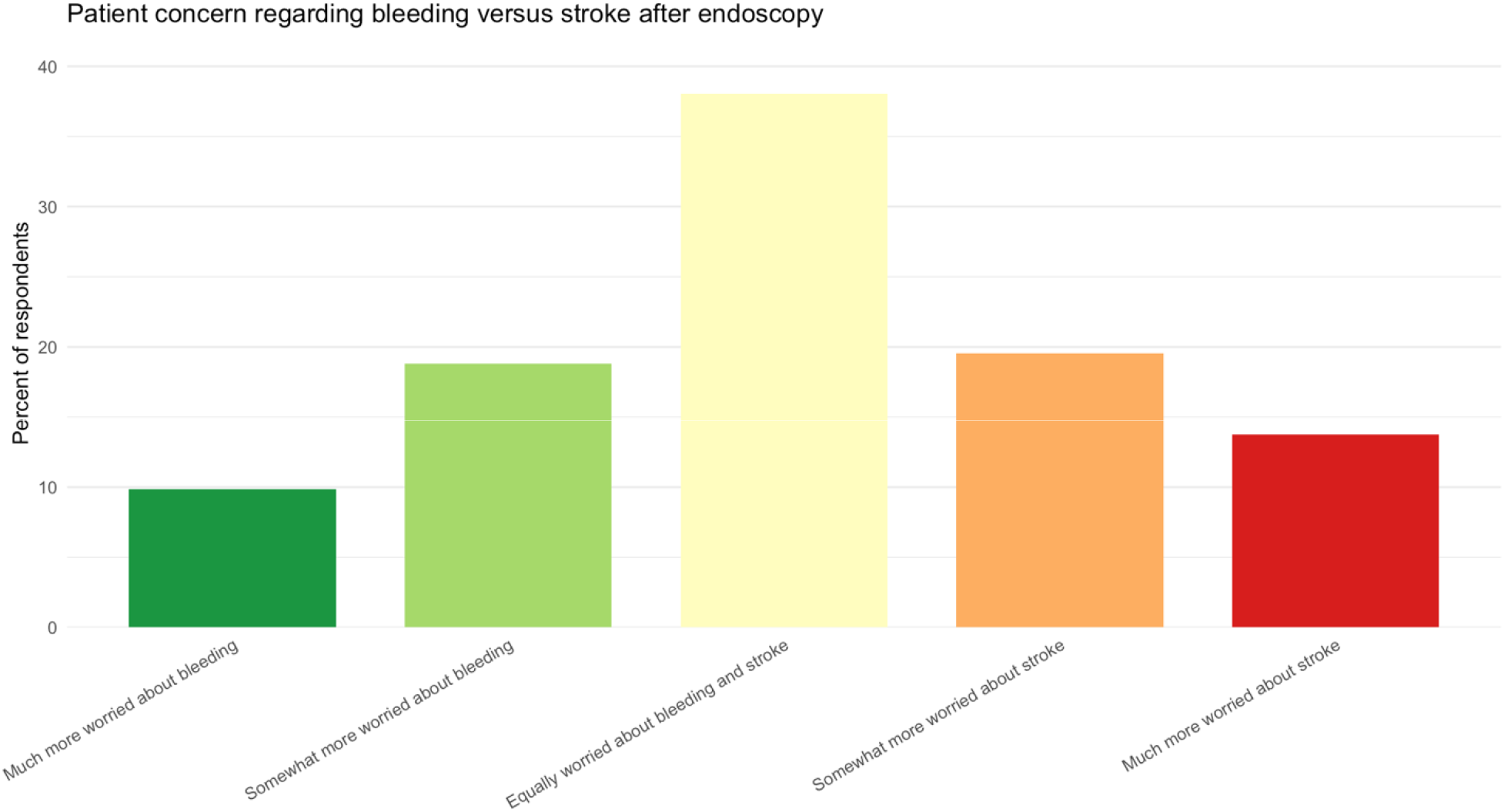
Ordinal response data from patients regarding what outcome of DOAC interruption is feared more (bleeding vs stroke).

### Influence of prior adverse clinical events on patient preferences

Exploratory subgroup analyses examined whether prior clinical events influenced patient outcome prioritization. Neither forced-choice responses nor ordinal preference scores differed significantly between respondents with and without a history of prior stroke or transient ischemic attack. Similarly, preferences did not differ between respondents with versus without a history of prior major bleeding **(Table 4)**, suggesting that prior adverse clinical experiences did not meaningfully shift patient prioritization of bleeding versus thromboembolic risk.

### Influence of demographic characteristics on patient preferences

Additional analyses assessed whether demographic characteristics were associated with ordinal outcome prioritization. Preferences did not differ significantly by age group, sex, or geographic region **(Supplemental Table 16)**. Across all demographic subgroups, the distribution of responses remained centered on the neutral response, with similar proportions of respondents expressing greater concern for bleeding, greater concern for stroke, or equal concern for both outcomes.

## Discussion

In this pair of surveys assessing both clinicians and patients, we identified substantial uncertainty and variability regarding the optimal timing of DOAC resumption following high-risk endoscopic procedures. Among endoscopists, practice patterns varied widely, and most respondents found more than one resumption strategy acceptable. Among patients, preferences regarding bleeding versus thromboembolic risk were heterogeneous and centered around the neutral response, without dominance of a single outcome priority. Together, these findings support the ethical justification, feasibility, and relevance of the proposed RESUME trial.

All current practice guidelines across the world provide concise recommendations regarding the time to withhold DOACs prior to elective GI endoscopy, which is critical for minimizing the risk of intraprocedural bleeding ^7-14^. While intraprocedural bleeding is important, it is delayed, post-procedure bleeding which is the more feared complication of GI endoscopy. Delayed bleeding typically results in hospital admission and often requires the transfusion of blood products and urgent repeat endoscopy to achieve hemostasis^20-22^. The management of delayed bleeding after GI endoscopy is estimated to cost $7,000-$12,000 US dollars per occurrence and therefore, is of significant interest not only to patients and family members, but also to healthcare systems and insurance payers^20, 22^. Studies have consistently shown that DOAC use is independently associated with higher rates of delayed bleeding after high-risk endoscopy ^23, 24^. Specifically, a large Canadian observational ERCP study showed that patients on DOAC therapy had a 7.3% risk of clinically significant delayed bleeding—a 4-fold increase compared to those that were not on DOACs^25^.

Although POD +2 was the most commonly selected resumption time in the survey vignette, RESUME is designed to compare strategy-based intervals that meaningfully differ in timing rather than adjacent single-day increments. Given the rapid onset of action of DOACs, a one-day difference in resumption may not translate into a clinically detectable difference in outcomes, particularly within the context of multifactorial bleeding risk after high-risk endoscopy. The selection of POD +1, +3, and +5 reflects a pragmatic approach intended to capture early, intermediate, and later resumption strategies that are already in use and that represent clinically distinguishable management philosophies. In this framework, POD +2 lies within the intermediate spectrum and is conceptually encompassed between the early and intermediate strategies. By comparing commonly employed resumption strategies, RESUME will fit well within a normal clinical workflow and is positioned to generate evidence that is directly applicable to routine clinical practice. This design reflects the reality that clinicians are already making these decisions daily in the absence of an evidence base to provide definitive guidance and that the resulting variability resumption timing, although modest, may have meaningful implications for both bleeding and thromboembolic outcomes.

This survey study has several strengths, including the inclusion of both clinician and patient perspectives, the use of complementary measures to assess preferences, and the consistency of findings across multiple subgroup analyses. However, limitations should be acknowledged. As a survey-based study, results may be influenced by selection bias, and respondents may not fully represent all endoscopists or patients receiving DOAC therapy. Patient respondents were recruited through advocacy organizations and may be more engaged than the general population. Additionally, responses were based on hypothetical scenarios, which may not fully capture real-world decision-making in acute clinical contexts. Nonetheless, the convergence of findings across stakeholders and the absence of strong subgroup effects support the robustness of the observed equipoise.

In conclusion, both clinicians and patients demonstrate substantial uncertainty and balanced preferences regarding the timing of DOAC resumption after high-risk endoscopic procedures. In the setting of limited guideline direction and widespread practice variability, these findings provide a strong rationale for a randomized evaluation of alternative resumption strategies for DOACs after high-risk endoscopic procedures. Addressing this evidence gap has the potential to inform future guidelines, reduce unwarranted practice variation, and improve patient-centered care in periprocedural anticoagulation management.

## Data Availability

All data produced in the present study are available upon reasonable request to the authors

## Abbreviations

DOAC: Direct Oral Anticoagulant
GI: Gastrointestinal
POD: Postoperative Day
EMR: Endoscopic Mucosal Resection
AF: Atrial Fibrillation
ACG: American College of Gastroenterology
CAG: Canadian Association of Gastroenterology
JGES: Japan Gastrointestinal Endoscopy Society
ASGE: American Society for Gastrointestinal Endoscopy
ERCP: Endoscopic Retrograde Cholangiopancreatography
ESD: Endoscopic Submucosal Dissection
US: United States
RESUME: Resumption of Direct Oral Anticoagulants After High-Risk Endoscopy
IRB: Institutional Review Board
MCW: Medical College of Wisconsin

## References

1. Barnes GD, Lucas E, Alexander GC, Goldberger ZD. National Trends in Ambulatory Oral Anticoagulant Use. Am J Med 2015;128:1300–5 e2.

2. Costa LSD, Alsultan MM, Hincapie AL, Guo JJ. Trends in utilization, reimbursement, and price for DOACs and warfarin in the US Medicaid population from 2000 to 2020. J Thromb Thrombolysis 2023;55:339–345.

3. Perreault S, de Denus S, White-Guay B, et al. Oral Anticoagulant Prescription Trends, Profile Use, and Determinants of Adherence in Patients with Atrial Fibrillation. Pharmacotherapy 2020;40:40–54.

4. Thyagaturu H, Seetharam K, Roma N, et al. Prescription Medication Use and Expenditure for Atrial Fibrillation in the United States. Value Health 2025;28:197–205.

5. Ho KH, van Hove M, Leng G. Trends in anticoagulant prescribing: a review of local policies in English primary care. BMC Health Serv Res 2020;20:279.

6. Kjerpeseth LJ, Ellekjaer H, Selmer R, et al. Trends in use of warfarin and direct oral anticoagulants in atrial fibrillation in Norway, 2010 to 2015. Eur J Clin Pharmacol 2017;73:1417–1425.

7. Abraham NS, Barkun AN, Sauer BG, et al. American College of Gastroenterology-Canadian Association of Gastroenterology Clinical Practice Guideline: Management of Anticoagulants and Antiplatelets During Acute Gastrointestinal Bleeding and the Periendoscopic Period. Am J Gastroenterol 2022;117:542–558.

8. Acosta RD, Abraham NS, Chandrasekhara V, et al. The management of antithrombotic agents for patients undergoing GI endoscopy. Gastrointest Endosc 2016;83:3–16.

9. Arora A, Kumar A, Anand AC, et al. Position statement from the Indian Society of Gastroenterology, Cardiological Society of India, Indian Academy of Neurology and Vascular Society of India on gastrointestinal bleeding and endoscopic procedures in patients on antiplatelet and/or anticoagulant therapy. Indian J Gastroenterol 2023;42:332–346.

10. Chan FKL, Goh KL, Reddy N, et al. Management of patients on antithrombotic agents undergoing emergency and elective endoscopy: joint Asian Pacific Association of Gastroenterology (APAGE) and Asian Pacific Society for Digestive Endoscopy (APSDE) practice guidelines. Gut 2018;67:405–417.

11. Cyrany J, Maly R, Rejchrt S, Tacheci I. Antithrombotic therapy and digestive endoscopy. Vnitr Lek 2022;68:538–542.

12. Kang SJ, Tae CH, Bang CS, et al. International Digestive Endoscopy Network consensus on the management of antithrombotic agents in patients undergoing gastrointestinal endoscopy. Clin Endosc 2024;57:141–157.

13. Kato M, Uedo N, Hokimoto S, et al. Guidelines for Gastroenterological Endoscopy in Patients Undergoing Antithrombotic Treatment: 2017 Appendix on Anticoagulants Including Direct Oral Anticoagulants. Dig Endosc 2018;30:433–440.

14. Veitch AM, Radaelli F, Alikhan R, et al. Endoscopy in patients on antiplatelet or anticoagulant therapy: British Society of Gastroenterology (BSG) and European Society of Gastrointestinal Endoscopy (ESGE) guideline update. Gut 2021;70:1611–1628.

15. Nagata N, Yasunaga H, Matsui H, et al. Therapeutic endoscopy-related GI bleeding and thromboembolic events in patients using warfarin or direct oral anticoagulants: results from a large nationwide database analysis. Gut 2018;67:1805–1812.

16. Radaelli F, Fuccio L, Paggi S, et al. Periendoscopic management of direct oral anticoagulants: a prospective cohort study. Gut 2019;68:969–976.

17. Rodriguez de Santiago E, Sanchez Aldehuelo R, Riu Pons F, et al. Endoscopy-Related Bleeding and Thromboembolic Events in Patients on Direct Oral Anticoagulants or Vitamin K Antagonists. Clin Gastroenterol Hepatol 2022;20:e380–e397.

18. Tomida H, Yoshio T, Igarashi K, et al. Influence of anticoagulants on the risk of delayed bleeding after gastric endoscopic submucosal dissection: a multicenter retrospective study. Gastric Cancer 2021;24:179–189.

19. Yanagisawa N, Nagata N, Watanabe K, et al. Post-polypectomy bleeding and thromboembolism risks associated with warfarin vs direct oral anticoagulants. World J Gastroenterol 2018;24:1540–1549.

20. Mehta D, Loutfy AH, Kushnir VM, et al. Cold versus hot endoscopic mucosal resection for large sessile colon polyps: a cost-effectiveness analysis. Endoscopy 2022;54:367–375.

21. Parikh ND, Zanocco K, Keswani RN, Gawron AJ. A cost-efficacy decision analysis of prophylactic clip placement after endoscopic removal of large polyps. Clin Gastroenterol Hepatol 2013;11:1319–24.

22. Shah ED, Pohl H, Rex DK, et al. Routine Prophylactic Clip Closure Is Cost Saving After Endoscopic Resection of Large Colon Polyps in a Medicare Population. Gastroenterology 2020;158:1164–1166 e3.

23. Masuda S, Koizumi K, Nishino T, et al. Direct oral anticoagulants increase bleeding risk after endoscopic sphincterotomy: a retrospective study. BMC Gastroenterology 2021;21:401.

24. Parras Castanera E, Rodriguez Lopez P, Alvarez A, et al. Predictive factors for post-ERCP bleeding. Influence of direct oral anticoagulants. Revista Espanola de Enfermedades Digestivas 2021;113:591–596.

25. Bishay K, Ruan Y, Barkun AN, et al. Incidence, Predictors, and Outcomes of Clinically Significant Post-Endoscopic Retrograde Cholangiopancreatography Bleeding: A Contemporary Multicenter Study. Am J Gastroenterol 2024;119:2317–2325.

