## Supplemental Material for "Clinical equipoise and patient preferences for DOAC resumption after high-risk endoscopy: implications for a randomized trial"

Clinical equipoise and patient preferences for direct oral anticoagulant resumption after high-risk endoscopy: implications for a randomized trial design

Supplemental Material

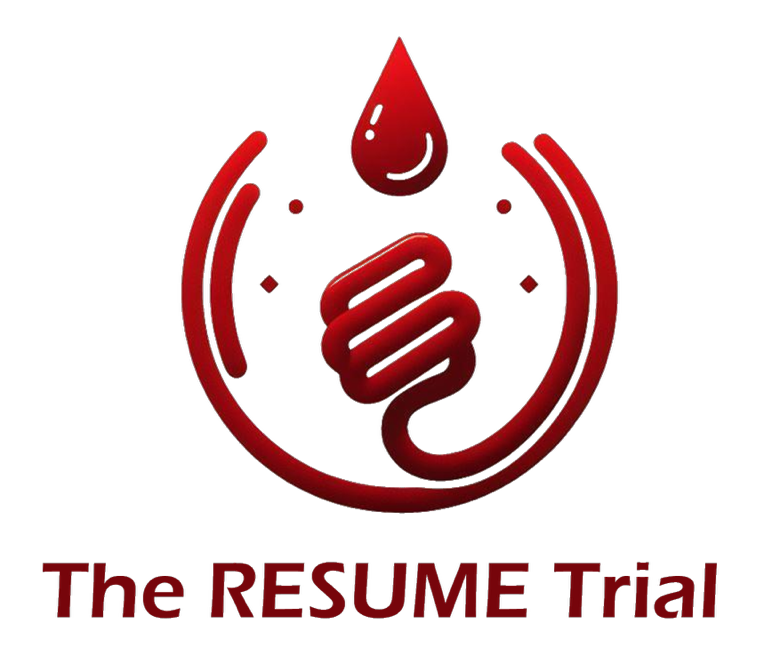

### Endoscopist Survey

RESUME Letter of Request ASGE Survey

Start of Block: Introduction

Q1 The **RESUME Trial** is a proposed NIH-funded, multicenter randomized trial evaluating the **optimal timing of direct oral anticoagulant (DOAC) resumption after high-risk endoscopic procedures**. Delayed bleeding after procedures such as EMR, ESD, POEM, or sphincterotomy is a serious clinical concern, especially in patients on DOACs. However, there is no robust evidence to guide when anticoagulation should be restarted, and current decisions are largely based on personal experience or local practice. RESUME is designed to compare three post-procedure DOAC resumption strategies. Preliminarily, these resumption strategies are **early** (postoperative day (POD) 1), **intermediate** (POD 3), or **late** (POD 5). The goal of RESUME is to generate definitive guidance for this common and important clinical scenario. To inform the design of RESUME—centered around **stakeholder engagement of endoscopists** of varying experience and practice settings—we hope to design a trial rooted in equipoise and to generate results that are generalizable to a wide range of clinicians. The following survey should take **no more than 5 minutes** to complete, and its results will be highly valuable in helping inform the design of RESUME for a competitive funding application. We thank you in advance for your consideration and time in participating in what we collectively feel is a critically important clinical trial.

**Zachary L. Smith, DO, MSc.**

Medical College of Wisconsin and Wisconsin Institute for Research in Endoscopy

On behalf of the **RESUME Trial Investigators**

| Page Break |
| --- |

Hypothetical Case  You perform a conventional (i.e. “hot”) endoscopic mucosal resection (EMR) of a 35 mm sessile polyp in the ascending colon in a 74-year-old patient taking apixaban for non-valvular atrial fibrillation. The lesion is removed successfully, but the site cannot be clipped closed due to its location. In the absence of additional patient-specific risk factors (e.g. prior stroke or GI bleed), **how many days post-procedure would you typically wait before resuming apixaban** (choose the single best answer)?

- 0 days (resume same day) (1)
- 1 day (2)
- 2 days (3)
- 3 days (4)
- 4 days (5)
- 5 days (6)
- More than 5 days (7)
- I am unsure (8)
- Defer to cardiology / prescribing physician (9)
- Other (10)

| Page Break |
| --- |

Equipoise Assessment

The RESUME Trial will compare three DOAC resumption strategies after high-risk endoscopy. **Preliminarily**, these strategies are:  •	**Early resumption**: Postoperative day 1 (POD+1)  •	**Intermediate resumption**: Postoperative day 3 (POD+3)  •	**Late resumption**: Postoperative day 5 (POD+5) These reflect the variation currently seen in clinical practice. We’d like to know your perspective on the acceptability of each strategy in the preliminary trial design. In a high-risk endoscopic procedure setting—such as the prior hypothetical case—how acceptable is each of the following DOAC resumption strategies from a clinical standpoint?

|  | Not Acceptable (1) | Acceptable, with reservations (2) | Acceptable (3) | Strongly prefer (4) |
| --- | --- | --- | --- | --- |
| Postoperative Day 1 (early) (1) |  |  |  |  |
| Postoperative Day 3 (intermediate) (2) |  |  |  |  |
| Postoperative Day 5 (late) (3) |  |  |  |  |

| Page Break |
| --- |

Value of Study

How important is it to have a well-powered randomized trial to guide DOAC resumption timing after high-risk endoscopy?

- Not at all important (1)
- Slightly important (2)
- Moderately important (3)
- Very important (4)
- Extremely important (5)

| Page Break |
| --- |

Additional Arms

In addition to postoperative day (POD) 1, 3, and 5, which of the following DOAC resumption strategies do you think would be worth evaluating in a clinical trial? (Select all that apply)

- POD 0 (same-day resumption) (1)
- POD 2 (2)
- POD 4 (3)
- POD 6-7 (one week) (4)
- Individualized based on bleeding risk factors (5)
- Individualized based on thromboembolic risk (6)
- Shared decision-making / patient preference (7)
- I do not believe other strategies need to be studied (8)
- Other (9) __________________________________________________

| Page Break |
| --- |

Q13

When deciding when to restart a DOAC after a high-risk gastrointestinal procedure (such as the prior hypothetical EMR case), doctors must balance the risks of bleeding and stroke. Below are several possible outcomes that could occur after such a procedure. **Please rank these outcomes from most serious (1) to least serious (6)** based on **what would matter most to you as the performing endoscopist**.

______ Delayed GI bleeding that requires blood transfusion, ICU care, or another urgent procedure (such as colonoscopy, angiography, or surgery) (1)

______ Delayed GI bleeding that resolves on its own but requires hospitalization for observation or monitoring (2)

______ Transient ischemic attack (TIA) – a “mini-stroke” with complete recovery of neurologic symptoms (3)

______ Stroke with long-term or permanent neurologic symptoms (e.g., weakness, speech difficulty, or paralysis) (4)

______ Death from delayed GI bleeding (5)

______ Death from stroke (6)

End of Block: Introduction

Start of Block: Respondent Demographics

Q7 What is your age?

- Under 35 (1)
- 35-44 (2)
- 45-54 (3)
- 55-64 (4)
- 65 or older (5)
- Prefer not to answer (6)

Gender What is your gender?

- Male (1)
- Female (2)
- Non-binary / third gender (3)
- Prefer not to say (4)

Experience How many years have you been in independent clinical practice (following your last year of fellowship training)?

- 0-5 years (1)
- 6-10 years (2)
- 11-15 years (3)
- 16-20 years (4)
- 21 or more years (5)
- Prefer not to answer (6)

Region In which global region do you primarily practice?

- US - Northeast (1)
- US - Mid-Atlantic (2)
- US - Southeast (3)
- US - Central Plains (4)
- US - Midwest (5)
- US - Mountain West (6)
- US - Southwest (7)
- US - West Coast (8)
- Canada (9)
- Mexico / Latin America (10)
- South America (11)
- Ireland / United Kingdom (12)
- Western Europe (13)
- Eastern Europe (14)
- Asia - Western (15)
- Asia - South (16)
- Asia - Southeast (17)
- Asia - Central (18)
- Asia - East (19)
- Oceania (20)
- Africa (21)
- Other (22)

Practice What is your primary practice setting?

- Academic medical center (1)
- Private practice (2)
- Hybrid academic-community practice (3)
- Veterans Affairs / Government / Military (4)
- Other (5)
- Prefer not to answer (6)

ERCPs How many ERCPs do you perform per year (on average)?

- 0-25 (1)
- 26-50 (2)
- 51-100 (3)
- 101-200 (4)
- More than 200 (5)
- I do not perform ERCP (6)
- Prefer not to answer (7)

Polyp resection How many large polyp resections (≥20 mm EMR/ESD) do you perform per year (on average)?

- 0-10 (1)
- 11-25 (2)
- 26-50 (3)
- 51-99 (4)
- 100 or more (5)
- I do not do large colorectal polyp resection (6)
- Prefer not to answer (7)

End of Block: Respondent Demographics

### Patient Survey

RESUME Patient Survey

Start of Block: Default Question Block

Introduction You are invited to take part in a brief, anonymous survey being conducted by researchers at the **Medical College of Wisconsin**. Our team is preparing a large research grant application to the **National Institutes of Health (NIH)** to **study how to safely restart direct oral anticoagulant (DOAC) medications (e.g. apixaban, rivaroxaban, edoxaban, dabigatran) after certain high-risk gastrointestinal (GI) procedures**. To make sure this study reflects what matters most to patients, **we are asking for your input** on how you view different strategies for stopping and restarting these medications. Your responses will help guide the design of a major national clinical trial aimed at improving the safety and quality of care for patients who take anticoagulants. This survey should take about **5 minutes** to complete. Participation is voluntary and anonymous. You may skip any question or stop at any time. On behalf of all my RESUME co-investigators, I thank you in advance for your participation. Sincerely, Zachary L. Smith, DO, MSc. Principal Investigator, RESUME Trial Associate Professor of Medicine Medical College of Wisconsin

| Page Break |
| --- |

End of Block: Default Question Block

Start of Block: Section 1: About You

Q1 What is your age group?

- Under 40 (1)
- 40-54 (2)
- 55-64 (3)
- 65-74 (4)
- 75 or over (5)
- Prefer not to answer (6)

Q2 What is your gender?

- Male (1)
- Female (2)
- Non-binary / third gender (3)
- Prefer not to say (4)

Q3 What continent do you live on?

- North America (1)
- Europe (2)
- South America (3)
- Asia (4)
- Australia (5)
- Africa (6)

Q4 Are you currently taking an anticoagulant (**NOT including warfarin/Coumadin**) such as Eliquis (apixaban), Xarelto (rivaroxaban), Pradaxa (dabigatran), or Savaysa (edoxaban)?

- Yes (1)
- No (2)
- I have taken one in the past but not currently (3)
- Unsure (4)
- Prefer not to answer (5)

Q5 Have you ever experienced a neurologic event such as a stroke or TIA (“mini-stroke”)?

- Yes (1)
- No (2)
- I am not sure (3)

Q6 Have you ever experienced a **major** bleeding event, such as bleeding from the stomach or intestines (GI bleeding) or another major bleed?

- Yes (1)
- No (2)
- I am not sure (3)

Q7 Have you ever had a gastrointestinal (GI) procedure such as a colonoscopy or endoscopy?

- Yes (1)
- No (2)
- I am not sure (3)

End of Block: Section 1: About You

Start of Block: Awareness and Understanding

Q8 How confident are you that there is **clear scientific evidence** or medical guidance on **when to restart anticoagulants after procedures that are high risk for delayed bleeding** (such as colonoscopy with removal of a large polyp)?

- Very confident that clear guidance exists (1)
- Somewhat confident that clear guidance exists (2)
- Unsure (3)
- Not very confident that clear guidance exists (4)
- There is no clear guidance on this topic (5)

End of Block: Awareness and Understanding

Start of Block: Importance of Different Possible Outcomes

Q9 When thinking about restarting an anticoagulant after a high-risk procedure, which of the following concerns you the MOST? (Select one)

- Major bleeding that requires hospitalization or an urgent procedure (1)
- Stroke that causes long-term disability (3)
- I'm not sure (5)

Q10 Which risk worries you more when restarting your anticoagulant after a procedure?

- Much more worried about bleeding (1)
- Somewhat more worried about bleeding (2)
- Equally worried about bleeding and stroke (3)
- Somewhat more worried about stroke (4)
- Much more worried about stroke (5)
- I'm not sure (6)

End of Block: Importance of Different Possible Outcomes

### Supplemental Table 1: Perceived importance of a randomized trial examining DOAC resumption after high-risk endoscopy.

How important is it to have a well-powered randomized trial to guide DOAC resumption timing after high-risk endoscopy?

|  | n (%) |
| --- | --- |
| Not at all important | 2 (1.1%) |
| Slightly important | 5 (2.8%) |
| Moderately important | 26 (14.7%) |
| Very important | 80 (45.2%) |
| Extremely important | 64 (36.2%) |
| Total | **177 (100%)** |

### Supplemental Table 2: Hypothetical case

You perform a conventional (i.e. “hot”) endoscopic mucosal resection (EMR) of a 35 mm sessile polyp in the ascending colon in a 74-year-old patient taking apixaban for non-valvular atrial fibrillation. The lesion is removed successfully, but the site cannot be clipped closed due to its location. In the absence of additional patient-specific risk factors (e.g. prior stroke or GI bleed), how many days post-procedure would you typically wait before resuming apixaban (choose the single best answer)?

|  | n (%) |
| --- | --- |
| 0 days (resume same day) | 8 (4.3%) |
| 1 day | 33 (17.8%) |
| 2 days | 68 (36.8%) |
| 3 days | 34 (18.4%) |
| 4 days | 4 (2.2%) |
| 5 days | 26 (14.1%) |
| More than 5 days | 5 (2.7%) |
| I am unsure | 2 (1.1%) |
| Defer to cardiology / prescribing physician | 5 (2.7%) |
| Total | **185 (100%)** |

### Supplemental Table 3: Assessment of Equipoise

The RESUME Trial will compare three DOAC resumption strategies after high-risk endoscopy. Preliminarily, these strategies are: • Early resumption: Postoperative day 1 (POD+1) • Intermediate resumption: Postoperative day 3 (POD+3) • Late resumption: Postoperative day 5 (POD+5). These reflect the variation currently seen in clinical practice. We’d like to know your perspective on the acceptability of each strategy in the preliminary trial design.

In a high-risk endoscopic procedure setting—such as the prior hypothetical case—how acceptable is each of the following DOAC resumption strategies from a clinical standpoint?

|  | POD+1 (early) | POD+3 (intermediate) | POD+5 (late) |
| --- | --- | --- | --- |
| Not Acceptable | 49 (27.7%) | 9 (5.5%) | 48 (29.3%) |
| Not Acceptable / Acceptable, with reservations | 0 (0%) | 0 (0%) | 1 (0.6%) |
| Acceptable, with reservations | 78 (44.1%) | 41 (24.8%) | 47 (28.7%) |
| Acceptable, with reservations / Acceptable | 0 (0%) | 1 (0.6%) | 0 (0%) |
| Acceptable | 39 (22%) | 92 (55.8%) | 48 (29.3%) |
| Acceptable / Strongly prefer | 3 (1.7%) | 1 (0.6%) | 0 (0%) |
| Strongly prefer | 8 (4.5%) | 21 (12.7%) | 20 (12.2%) |
| Total | **177 (100%)** | **165 (100%)** | **164 (100%)** |

### Supplemental Table 4: Additional Arms

In addition to postoperative day (POD) 1, 3, and 5, which of the following DOAC resumption strategies do you think would be worth evaluating in a clinical trial? (Select all that apply)

A total of 176 individuals provided a response to this question.

|  | n(%) |
| --- | --- |
| POD 0 (same-day resumption) | 55 (31.2) |
| POD 2 | 55 (31.2) |
| POD 4 | 9 (5.1) |
| POD 6-7 (one week) | 33 (18.8) |
| Individualized based on bleeding risk factors | 87 (49.4) |
| Individualized based on thromboembolic risk | 112 (63.6) |
| Shared decision-making / patient preference | 37 (21) |
| I do not believe other strategies need to be studied | 14 (8) |
| Other | 6 (3.4) |

### Supplemental Table 5: Ranked Outcomes (Endoscopist)

When deciding when to restart a DOAC after a high-risk gastrointestinal procedure (such as the prior hypothetical EMR case), doctors must balance the risks of bleeding and stroke. Below are several possible outcomes that could occur after such a procedure. Please rank these outcomes from most serious (1) to least serious (6) based on what would matter most to you as the performing endoscopist.

A total of 120 individuals provided a response to this question.

|  | min | 25 %tile | median | 75 %tile | max |
| --- | --- | --- | --- | --- | --- |
| Delayed GI bleeding that requires blood transfusion, ICU care, or another urgent procedure (such as colonoscopy, angiography, or surgery) | 1 | 4.0 | 4 | 5 | 6 |
| Delayed GI bleeding that resolves on its own but requires hospitalization for observation or monitoring | 1 | 5.0 | 6 | 6 | 6 |
| Transient ischemic attack (TIA) – a “mini-stroke” with complete recovery of neurologic symptoms | 1 | 4.0 | 5 | 5 | 6 |
| Stroke with long-term or permanent neurologic symptoms (e.g., weakness, speech difficulty, or paralysis) | 1 | 3.0 | 3 | 3 | 6 |
| Death from delayed GI bleeding | 1 | 1.5 | 2 | 2 | 6 |
| Death from stroke | 1 | 1.0 | 1 | 2 | 6 |

### Supplemental Table 6: Association between endoscopist experience (years in practice) and opinion of POD+1 as a trial arm.

|  | **POD + 1** | | **Total** | p-value*^1^* |
| --- | --- | --- | --- | --- |
|  | Not Acceptable | Acceptable |  |  |
| Experience |  |  |  | 0.5 |
| 0-10 years | 18 (25%) | 53 (75%) | 71 (100%) |  |
| 11+ years | 29 (30%) | 69 (70%) | 98 (100%) |  |
| Total | 47 (28%) | 122 (72%) | 169 (100%) |  |
| *^1^* Pearson’s Chi-squared test | | | | |

### Supplemental Table 7: Association between endoscopist practice setting and opinion of POD+1 as a trial arm.

|  | **POD + 1** | | **Total** | p-value*^1^* |
| --- | --- | --- | --- | --- |
|  | Not Acceptable | Acceptable |  |  |
| Practice Setting |  |  |  | 0.9 |
| Academic medical center | 17 (28%) | 44 (72%) | 61 (100%) |  |
| Private practice | 17 (30%) | 40 (70%) | 57 (100%) |  |
| Hybrid + VA + Other | 12 (25%) | 36 (75%) | 48 (100%) |  |
| Total | 46 (28%) | 120 (72%) | 166 (100%) |  |
| *^1^* Pearson’s Chi-squared test | | | | |

### Supplemental Table 8: Association between ERCP volume and opinion of POD+1 as a trial arm.

|  | **POD + 1** | | **Total** | p-value*^1^* |
| --- | --- | --- | --- | --- |
|  | Not Acceptable | Acceptable |  |  |
| ERCPs |  |  |  | 0.043 |
| 0-25 / Do not perform | 20 (22%) | 73 (78%) | 93 (100%) |  |
| 26+ | 27 (36%) | 49 (64%) | 76 (100%) |  |
| Total | 47 (28%) | 122 (72%) | 169 (100%) |  |
| *^1^* Pearson’s Chi-squared test | | | | |

### Supplemental Table 9: Association between complex polyp resection volume and opinion of POD+1 as a trial arm.

|  | **POD + 1** | | **Total** | p-value*^1^* |
| --- | --- | --- | --- | --- |
|  | Not Acceptable | Acceptable |  |  |
| Polyp resection |  |  |  | 0.6 |
| 0-10 / Do not perform | 7 (22%) | 25 (78%) | 32 (100%) |  |
| 11+ | 29 (26%) | 81 (74%) | 110 (100%) |  |
| Total | 36 (25%) | 106 (75%) | 142 (100%) |  |
| *^1^* Pearson’s Chi-squared test | | | | |

### Supplemental Table 10: Association between endoscopist experience (years in practice) and opinion of POD+5 as a trial arm.

|  | **POD + 5** | | **Total** | p-value*^1^* |
| --- | --- | --- | --- | --- |
|  | Not Acceptable | Acceptable |  |  |
| Experience |  |  |  | >0.9 |
| 0-10 years | 20 (29%) | 48 (71%) | 68 (100%) |  |
| 11+ years | 27 (30%) | 64 (70%) | 91 (100%) |  |
| Total | 47 (30%) | 112 (70%) | 159 (100%) |  |
| *^1^* Pearson’s Chi-squared test | | | | |

### Supplemental Table 11: Association between endoscopist practice setting and opinion of POD+5 as a trial arm.

|  | **POD + 5** | | **Total** | p-value*^1^* |
| --- | --- | --- | --- | --- |
|  | Not Acceptable | Acceptable |  |  |
| Practice Setting |  |  |  | 0.7 |
| Academic medical center | 16 (28%) | 41 (72%) | 57 (100%) |  |
| Private practice | 14 (26%) | 39 (74%) | 53 (100%) |  |
| Hybrid + VA + Other | 15 (33%) | 30 (67%) | 45 (100%) |  |
| Total | 45 (29%) | 110 (71%) | 155 (100%) |  |
| *^1^* Pearson’s Chi-squared test | | | | |

### Supplemental Table 12: Association between ERCP volume and opinion of POD+5 as a trial arm.

|  | **POD + 5** | | **Total** | p-value*^1^* |
| --- | --- | --- | --- | --- |
|  | Not Acceptable | Acceptable |  |  |
| ERCPs |  |  |  | 0.7 |
| 0-25 / Do not perform | 25 (30%) | 57 (70%) | 82 (100%) |  |
| 26+ | 21 (28%) | 55 (72%) | 76 (100%) |  |
| Total | 46 (29%) | 112 (71%) | 158 (100%) |  |
| *^1^* Pearson’s Chi-squared test | | | | |

### Supplemental Table 13: Association between complex polyp resection volume and opinion of POD+5 as a trial arm.

|  | **POD + 5** | | **Total** | p-value*^1^* |
| --- | --- | --- | --- | --- |
|  | Not Acceptable | Acceptable |  |  |
| Polyp resection |  |  |  | 0.6 |
| 0-10 / Do not perform | 10 (33%) | 20 (67%) | 30 (100%) |  |
| 11+ | 30 (29%) | 74 (71%) | 104 (100%) |  |
| Total | 40 (30%) | 94 (70%) | 134 (100%) |  |
| *^1^* Pearson’s Chi-squared test | | | | |

### Supplemental Table 14: Patient perceptions of the level of evidence guiding the practice of DOAC resumption after high-risk endoscopy.

| Awareness of evidence gaps regarding anticoagulant resumption after endoscopy | |
| --- | --- |
|  | n (%) |
| Awareness of evidence gap (collapsed) | |
| Confident clear guidance exists | 259 (54.6%) |
| No clear guidance | 16 (3.4%) |
| Other / unclassified | 199 (42%) |
| Values are n (%), calculated within the analysis population. No imputation performed. | |

### Supplemental Table 15: Balance of fears for bleeding and stroke with DOAC interruption

| Distribution of patient preferences by outcome priority (Q10) | |
| --- | --- |
| Outcome priority | n (%) |
| Bleeding feared more (1–2) | 119 (28.7%) |
| Equal concern (3) | 158 (38.1%) |
| Stroke feared more (4–5) | 138 (33.3%) |
